# Exploring the associations between skeletal muscle echogenicity and physical function in aging adults: A systematic review with meta-analyses

**DOI:** 10.1101/2023.12.13.23299929

**Authors:** Dustin J Oranchuk, Stephan G Bodkin, Katie L Boncella, Michael O Harris-Love

**Affiliations:** Muscle Morphology, Mechanics, and Performance Laboratory, Department of Physical Medicine and Rehabilitation, University of Colorado Anschutz Medical Campus, Aurora, CO, United States; Department of Physical Therapy and Athletic Training, University of Utah, Salt Lake City, UT, United States

**Author notes:** **Corresponding author:** Dustin J Oranchuk, Denver, Colorado, United States of America. **Funding disclosure:** None. **Conflict of interest:** None.

**Keywords:** correlations, echo intensity, elderly, intramuscular fat, muscle health, musculoskeletal, strength

## Abstract

**Background:** Assessment and quantification of skeletal muscle within the aging population is vital for diagnosis, treatment, and injury/disease prevention. The clinical availability of assessing muscle quality through diagnostic ultrasound presents an opportunity to be utilized as a screening tool for function-limiting diseases. However, relationships between muscle echogenicity and clinical functional assessments require authoritative analysis. Thus, we aimed to 1) synthesize the literature to assess the relationships between skeletal muscle echogenicity and physical function in older (≥60 years) adults, 2) perform pooled analyses of relationships between skeletal muscle echogenicity and physical function, and 3) perform sub-analyses to determine between-muscle relationships.

**Methods:** CINAHL, Embase, MEDLINE, PubMed, and Web of Science databases were systematically searched to identify articles relating skeletal muscle echogenicity to physical function in older adults. Meta-analyses with and without sub-analysis for individual muscles were performed utilizing Fisher’s Z transformation for the most common measures of physical function. Fisher’s Z was back-transformed to Pearson’s *r* for interpretation.

**Results:** Fifty-one articles (N=5095, female=∼2759, male=∼2301, 72.5±5.8 years [one study did not provide sex descriptors]) were extracted for review, with previously unpublished data obtained from the authors of 12 studies. The rectus femoris (n=32) and isometric knee extension strength (n=22) were the most accessed muscle and physical qualities, respectively. The relationship between quadriceps echogenicity and knee extensor strength was moderate (n=2924, *r*=-0.36 [95%CI: −0.38 to −0.32], *p*<0.001), with all other meta-analyses (grip strength, walking speed, sit-to-stand, timed up-and-go) resulting in slightly weaker correlations (*r*=−0.34 to −0.23, all *p*<0.001). Sub-analyses determined minimal differences in predictive ability between muscle groups, although combining muscles (e.g., rectus femoris+vastus lateralis) often resulted in stronger correlations with maximal strength.

**Conclusions:** While correlations were modest, the affordable, portable, and noninvasive ultrasonic assessment of muscle quality was a consistent predictor of physical function in older adults. Minimal between-muscle differences suggest that echogenicity estimates of muscle quality are systemic. Therefore, practitioners may be able to scan a single muscle to assess full-body skeletal muscle quality/composition, while researchers should consider combining multiple muscles to strengthen the model.

**Registration:** The original protocol was prospectively registered at the National Institute of Health Research PROSPERO (CRD42020201841).

**Highlights:**

- Relationships between skeletal muscle echogenicity and physical function were small to moderate, but highly consistent.
- Sub-analyses determined minimal between-muscle differences in predictive ability.
- Ultrasonic echogenicity should be considered part of early detection screens for sarcopenia and other diseases.
- Combining muscles tended to strengthen the model, although muscle quality appears systemic, allowing for a single scan to represent the total body.

## 1 Introduction

Assessment and quantification of skeletal muscle morphology and function within the aging population is vital for diagnosis, treatment, and injury/disease prevention. Sarcopenia, defined as the reduction in muscle mass and strength,^1^ is a growing concern, with up to 25% of individuals over 70 years receiving the diagnosis.^2^ Early identification of the decline in skeletal muscle morphology and function can lead to appropriate therapies, such as exercise or nutritional interventions, that may improve patient outcomes.^3^ To date, screening measures to identify patients at risk for sarcopenic-related disability include patient-reported outcomes (such as the SARC-F), objective measurement of grip or strength, and functional tasks such as the chair stand or 6-meter walk tests.^1,4^ In addition to the size and strength of skeletal muscle, recent literature has proposed the addition of skeletal muscle quality due to its relationship with patient outcomes and clinical accessibility.^5,6^

Muscle quality provides information on the morphology of the muscle, such as adipogenic, fibrotic, or contractile tissue.^7^ Literature has demonstrated increases in quadriceps intramuscular adipose and fibrous tissue with concurrent skeletal muscle atrophy with age, suggesting the potential replacement of contractile tissue.^8^ Fibrotic activity increases within aging muscle, which inhibits the ability to repair and regenerate.^9^ Clinical decisions made dependent solely on muscle size may fail to consider morphological characteristics of the muscle that impair physical function.

Changes in skeletal muscle morphology are traditionally assessed through invasive laboratory techniques – specifically by obtaining skeletal muscle biopsy to quantify the fibrotic or adipogenic tissue .^10^ Clinically accessible techniques to quantify intramuscular adipose tissue can be obtained through MRI and CT imaging. Though common, these techniques are time-consuming, cost-restrictive, and expose patients to radiation (CT scans). Diagnostic ultrasound quantifies muscle quality through echogenicity,^7^ which is defined as the pixel intensity of an image. Assessed through brightness mode (B-mode) ultrasound, the greyscale value (range: 0-255 arbitrary units represents the “quality” of the muscle, with a higher number representing a greater distribution of lighter pixels. A brighter pixel (higher greyscale value) represents lighter tissue, such as intramuscular adipocytes and fibrous tissue,^11^ while lower greyscale values have been related to greater lean body mass (e.g., contractile tissue, water).^11^

The clinical availability of assessing echogenicity also presents an opportunity to be utilized as a screening tool for function-limiting diseases including sarcopenia, cachexia, and muscular dystrophy. Lower muscle quality assessed through echogenicity has been associated with lower measures of muscle strength.^12^ However, results are not uniform within the aging population as assessments and techniques to quantify muscle function are inconsistent. Compiling research findings on the functional associates of echogenicity can further progress the clinical utility of ultrasound with aging individuals.

Early detection of muscle loss and strength is vital for early clinical diagnosis and treatment. Intramuscular adiposity has been shown to precede and accelerate the onset of sarcopenic functional impairments.^13^ As such, accessible clinical tools like diagnostic ultrasound may play an essential role in assessing muscle size and quality to predict loss of function. To date, relationships between muscle echogenicity and clinical and functional assessments are unclear due to the large variability in scanning location and assessment procedures. Therefore, the purpose of this systematic review was to 1) qualitatively synthesize the literature assessing the relationships between skeletal muscle echogenicity and physical function in older adults, 2) perform a quantitative pooled analyses of relevant correlations where possible, and 3) examine between-muscle relationships to determine differences in predictive ability.

## 2 Methods

### 2.1. Registration of systematic review protocol

A systematic literature review was performed according to the guidelines in the Cochrane Handbook for Systematic Reviews of Interventions (version 6.0) and following the 2020 checklist for the Preferred Reporting Items for Systematic Reviews and Meta-Analyses. The original protocol was prospectively registered at the National Institute of Health Research PROSPERO (CRD42020201841).

### 2.2. Eligibility criteria

Studies evaluating the relationship of ultrasound echogenicity to assessments of skeletal muscle function were included. All studies had to include an older population (≥60 years), quantified skeletal muscle echogenicity through diagnostic ultrasound, quantified clinical measures of patient function, and analyze the relationship between skeletal muscle echogenicity and patient function. For studies that included multiple age populations (i.e., a younger and older cohort,) only the data for the older cohort were extracted. Studies must have reported relationships between echogenicity and function performed through linear regression or correlation coefficients. Exclusion criteria included pathologic study populations, echogenicity outcomes on non-skeletal muscle tissue, animal or cadaver studies, or review articles. Studies including participants from a pathologic population (i.e., Parkinson’s, multiple sclerosis, muscular dystrophy, etc.) were excluded. Participants with age-related conditions, such as sarcopenia or osteoarthritis, were included.

### 2.3. Information sources and search strategy

Research articles were systematically searched through CINAHL, Embase, MEDLINE, PubMed, and Web of Science databases. Studies were searched up to June 2023. The current manuscript was a subsection of larger systematic search on relationships between diagnostic ultrasound echogenicity and metabolic, imaging, and functional outcomes (PROSPERO ID CRD42020201841). Search keywords were: (echogenicity OR echo intensity OR greyscale) AND (muscle OR musculature) AND (ultrasound OR ultrasonography) AND (old OR older OR elderly). The current studies’ outcomes of interest were relationships between skeletal muscle echogenicity and objective functional assessments (i.e., strength, gait, functional testing, patient-reported outcomes, etc.). Secondary searches included: a) screening the reference lists of included studies; b) examining studies that cited the included studies (forward citation tracking through Google Scholar); c) search alerts to monitor any new search results; and d) contacting the most common authors of the included outputs. The database search was re-run in November 2023.

### 2.4. Study appraisal and synthesis methods

Database results were downloaded and transferred to the Zotero reference manager (v6.0; Corporation for Digital Scholarship, Virginia, USA). Covidence (v2627; Melbourne, AUS) was used to import all selected articles from the initial search. Duplicate articles were removed for appraisal. Two authors independently screened the articles by titles and abstracts. The two reviewers met and discussed disagreements of the initial review to determine inclusion, with a third reviewer being consulted if needed. Both assessors then independently screened the full-text studies based on the inclusion and exclusion criteria using the same methodologies for disagreements. The PRISMA flowchart of article inclusion can be found in **Figure 1**. Data were extracted and were stratified on the following functional assessment categories: strength, rate of force development, gait speed, timed up-and-go, sit-to-stand, postural stability, and others. The reviewers assessed outcome measures to determine if inter-study data could be pooled for a meta-analysis.

**Figure 1.**
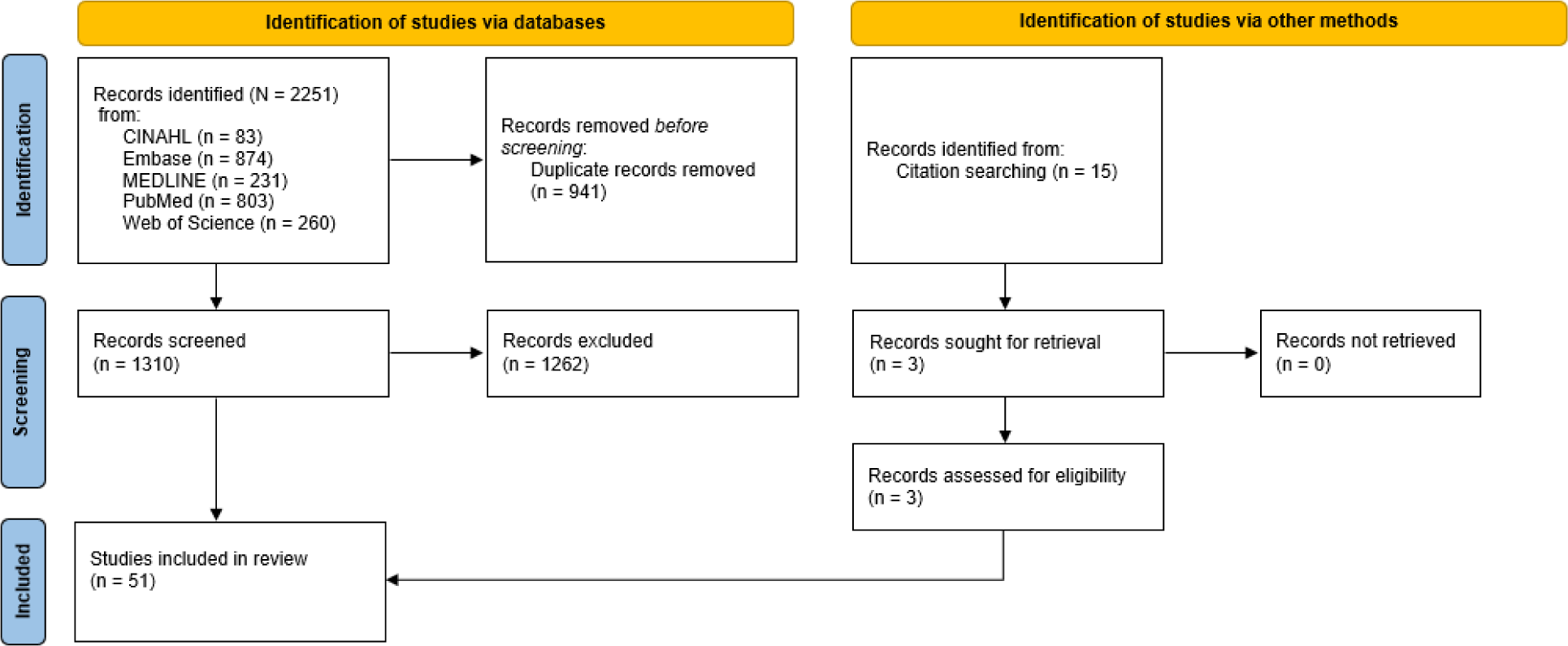
PRISMA flow chart.

### 2.5. Data extraction

Functional assessments, methods of collection and objective measures of the tests were extracted from each selected article. The transducer type, scanning plane, and muscle of interest were extracted for measures of muscle quality. Demographics, sample size, and relationship estimates (Pearson’s *r* correlation coefficients) were extracted for data synthesis.

### 2.6. Statistical analysis

#### 2.6.1. Correlation extraction and calculation

Raw data, reported correlation coefficients, and *p*-values were extracted and entered into an Excel spreadsheet. When correlations were not provided in-text, extracted data were used to calculate Pearson’s correlation coefficients (*r*) using the following formula (*r*=Pearson’s correlation, n=number of pairs, ∑xy = sum of the pairs, ∑x = sum of the x scores, ∑y= sum of the y scores, ∑x2 = sum of the squared x scores, ∑y2 = sum of the squared y scores:

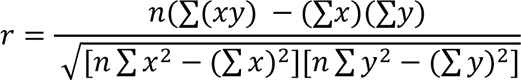

However, the above formula (and online calculators) often resulted in exceptionally strong correlations. Therefore, calculated correlations were not included in the meta-analyses. In several cases,^14–24^ contacted authors kindly provided de-identified raw data which allowed us to calculate exact correlations using SPSS (Version 29.0, IBM Corp; Armonk, NY, USA).

#### 2.6.2. Meta-analytical synthesis

As Pearson’s *r* correlation coefficients are a non-continuous variable (can never be greater than 1.0), all extracted and calculated correlations and standard errors were transformed to Fisher’s Z so that confidence intervals could be properly processed by the meta-analytical software. This transformation was completed using the following formula (Z=Fisher’s z, LN=natural log, r=Pearson’s *r* correlation coefficient:

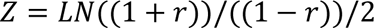

A random-effects model with restricted maximum likelihood was chosen due to inter-study variability regarding age, sex, racial/ethnic composition, and testing procedures. Pooled Fisher’s Z and 95% confidence intervals (95%CI) were back-transformed to Pearson’s *r* correlation coefficients and interpreted as: trivial ±0.10, small ±0.11−0.30, moderate ±0.31−0.50, large ±0.51−0.70, very large ±0.71−0.90, and nearly perfect ±0.91−0.99.^25^ Including multiple effects from a single study violates the assumption of independence, as effects from the same study are likely to be more similar than effects from others and would influence statistical power. Therefore, effects from a single study examining several regions of the same muscle (e.g., vastus lateralis, rectus femoris), or both sexes were averaged and entered the statistical software as a single comparison. Sub-analyses were run to compare different muscles. To avoid superfluous results, meta-analyses focused on muscle groups and highly relevant functional assessments (e.g., knee extensor strength and quadriceps echogenicity). However, several commonly reported functional assessments were not accompanied by clearly relevant echogenicity measures (e.g., grip strength and wrist flexor echogenicity). Similarly, we avoided performing meta-analyses with fewer than five studies. Finally, only correlations with non-corrected echogenicity were analyzed since most studies did not utilize subcutaneous fat correction. Similar tests (i.e., time to target repetitions versus repetitions in a set time) can result in dichotomous (i.e., positive versus negative) correlational directions. Thus, the correlation direction was switched when required to represent ‘performance’.

#### 2.6.3. Meta-analytical heterogeneity

The I^2^ statistic was used to evaluate heterogeneity and was interpreted as low (<50%), moderate (50–75%), and high heterogeneity (>75%). The statistical significance threshold was set at *p*<0.05. Funnel plots were used to examine publication bias potential.

## 3 Results

### 3.1. Study characteristics

The summary of studies selected for inclusion and their quality assessments are presented in Table 1. Fifty-one identified studies conformed to the inclusion criteria.^7,11,12,14–24,26–62^ However, 17 studies did not report the relevant correlations.^14–18,20,22,24,26,40,48,49,52,57,59,61,62^ 12 contacted authors provided previously unpublished de-identified raw data or relevant correlations.^14–20,22–24,40,48^ In comparison, we calculated correlations for the remaining five studies using data supplied in published tables or supplementary information. The 51 included studies included 5095 (∼2759 females, ∼2301 males) participants, aged 72.5±5.8 years of age.^7,11,12,14–24,26–62^ Twenty-seven studies included males and females,^14,17,20,23,24,26,28–30,32,33,36–38,40,42,45–47,49,54,57–62^ while 16 included only males,^7,11,15,16,18,19,21,22,31,34,39,41,48,50,52,56^ seven included only females,^27,35,43,44,51,53,55^ a single study did not report sex,^12^ while another did not delineate sex when reporting age or participant numbers;^61^ explaining the approximate values in the previous sentence. Two studies included participants with ‘metabolic syndrome’ or diabetes,^20,33^ and single studies included participants with chronic obstructive pulmonary disease,^42^ USA Veterans,^11^ ‘frail’,^24^ outpatients,^63^ nursing home residents,^23^ or regularly participated in competitive tennis.^59^ mmary of studies reporting correlations between skeletal muscle echogenicity and functional measures in older (Mean ≥ 63 years) adults

**Table 1.**
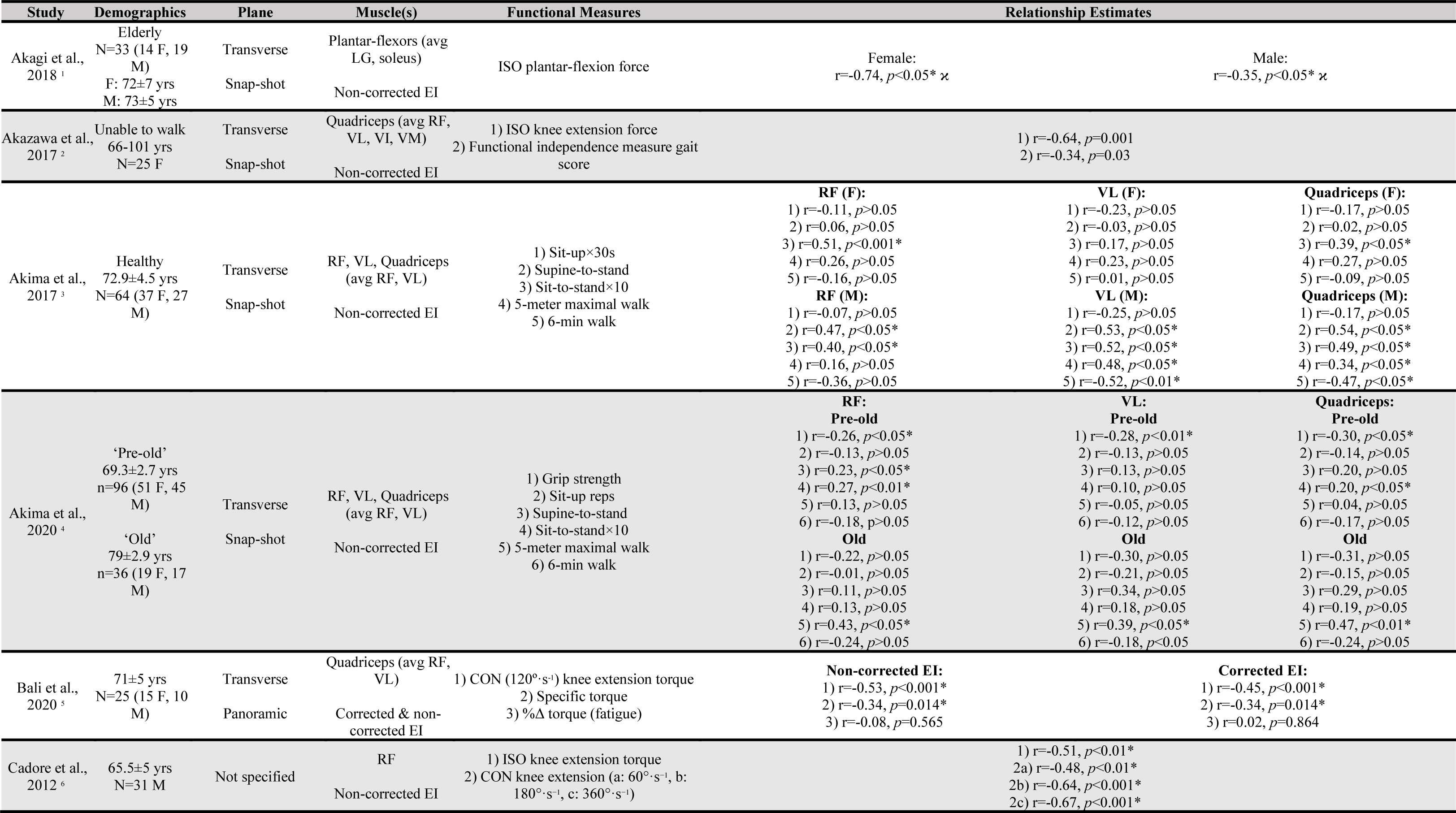
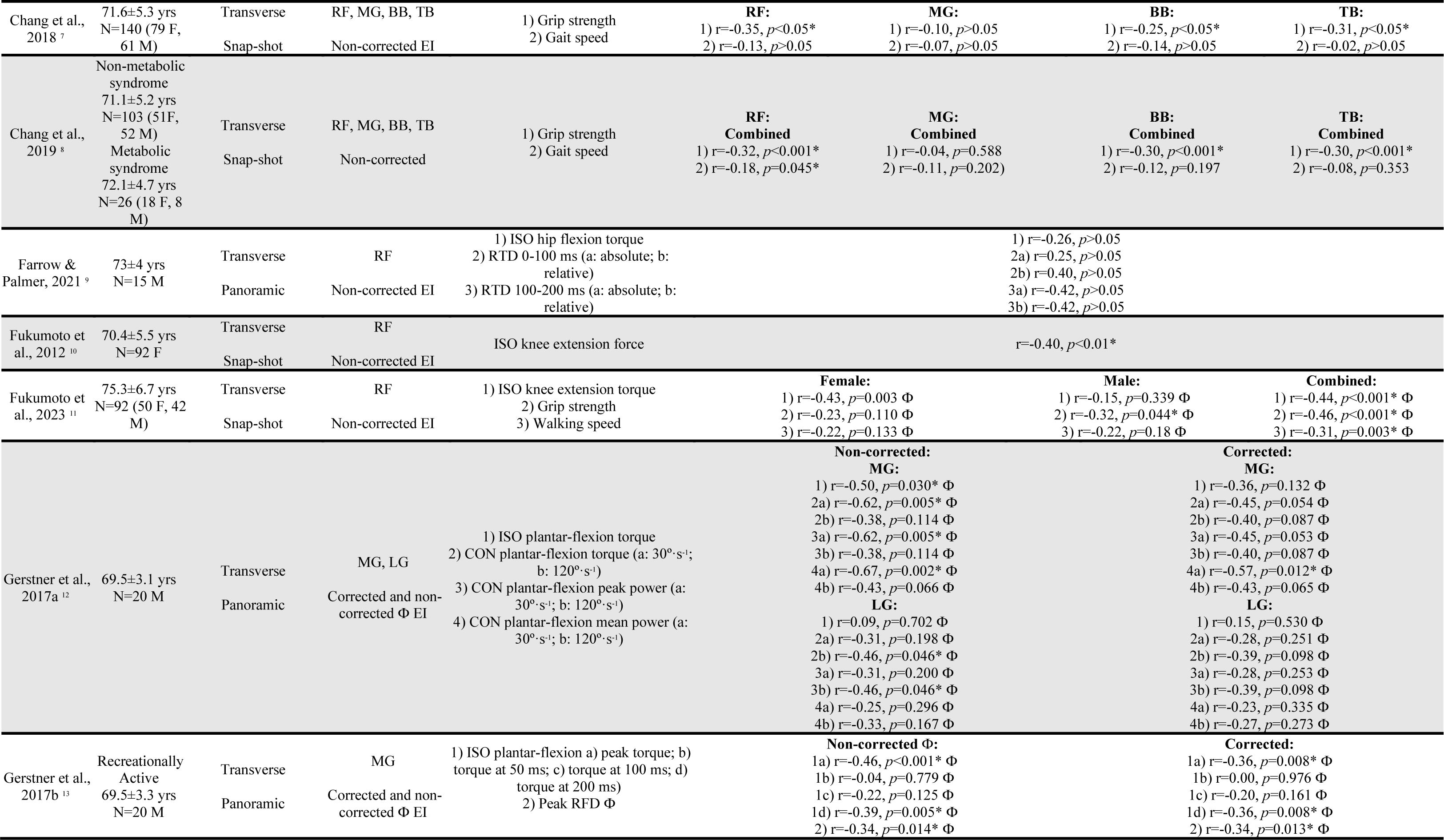
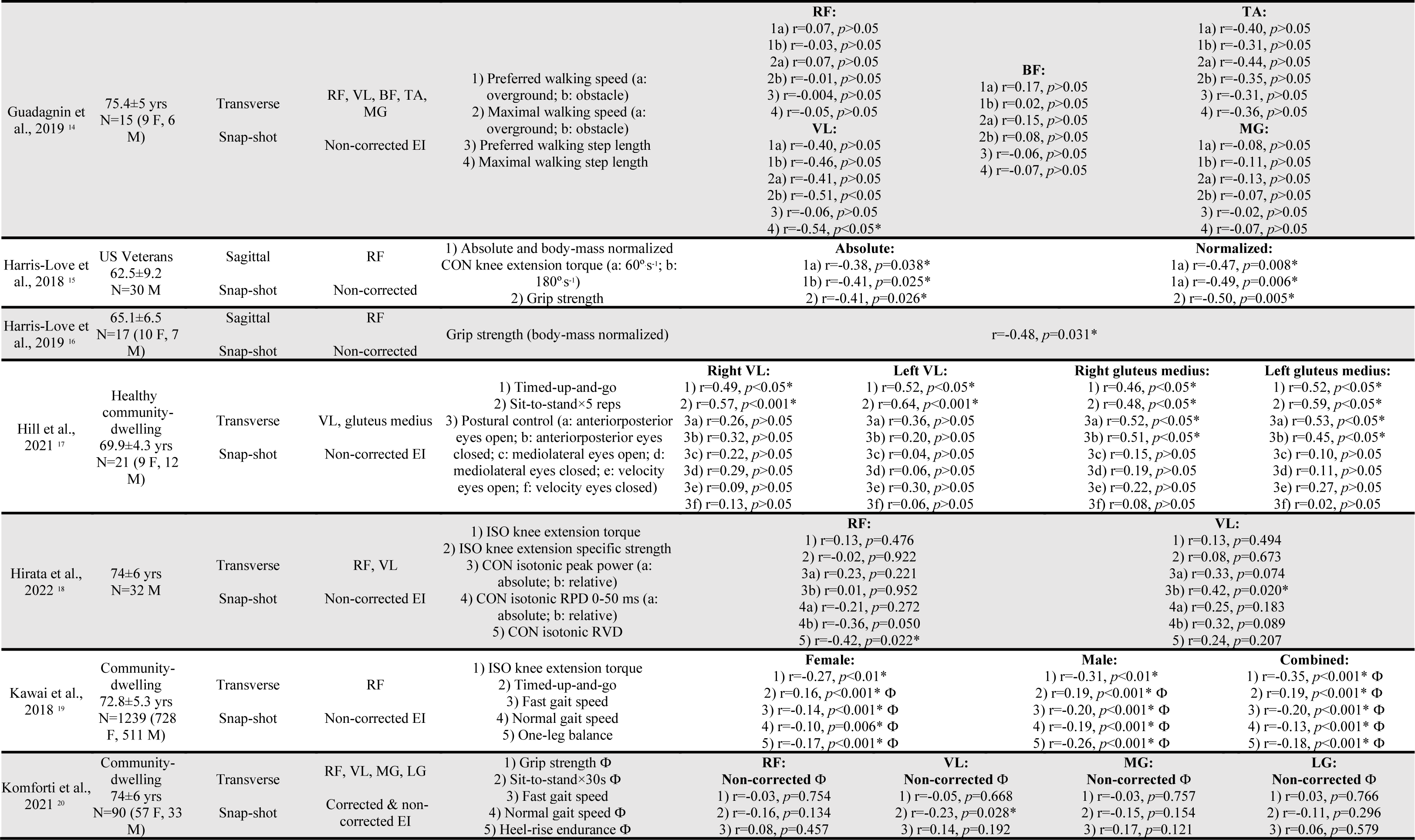
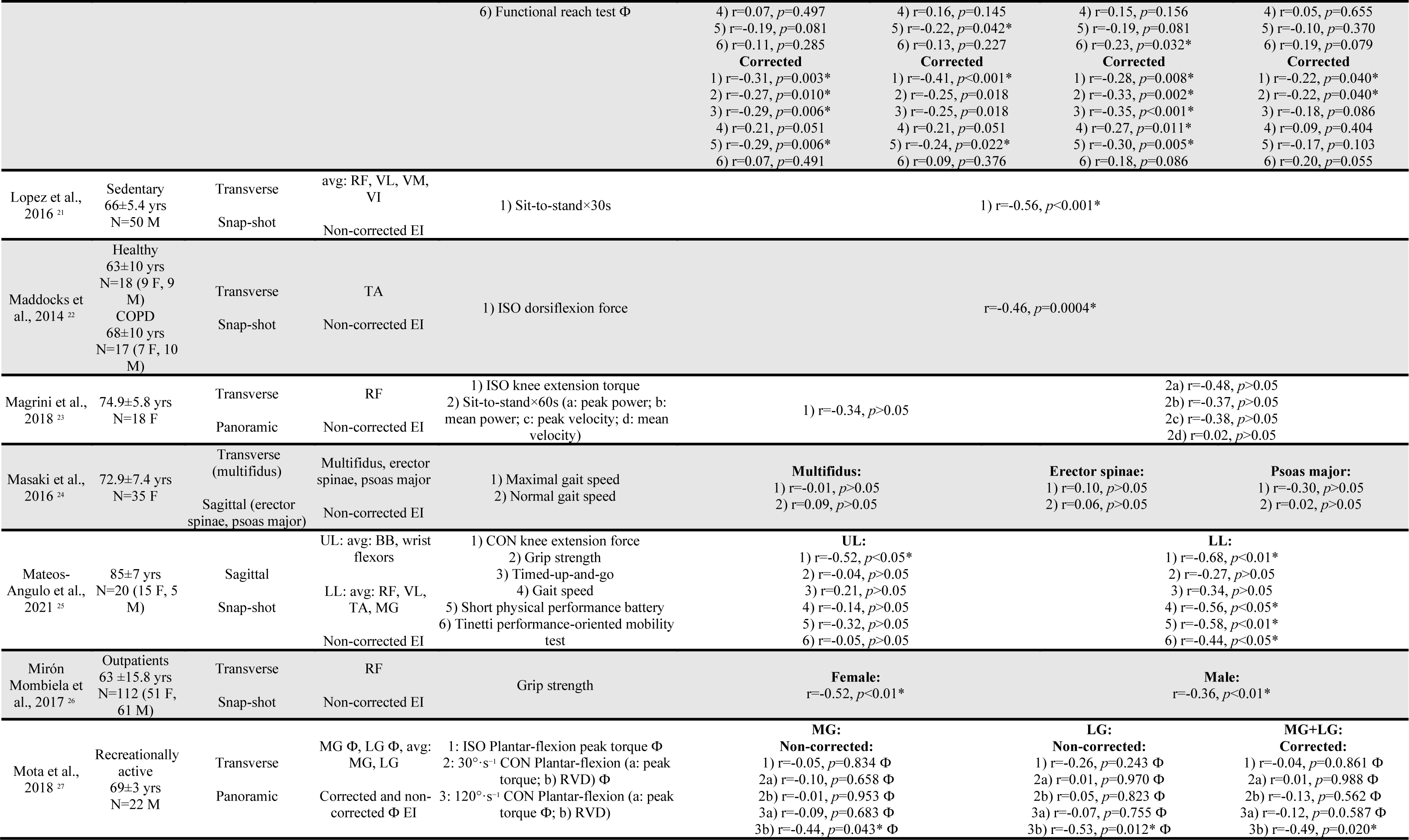
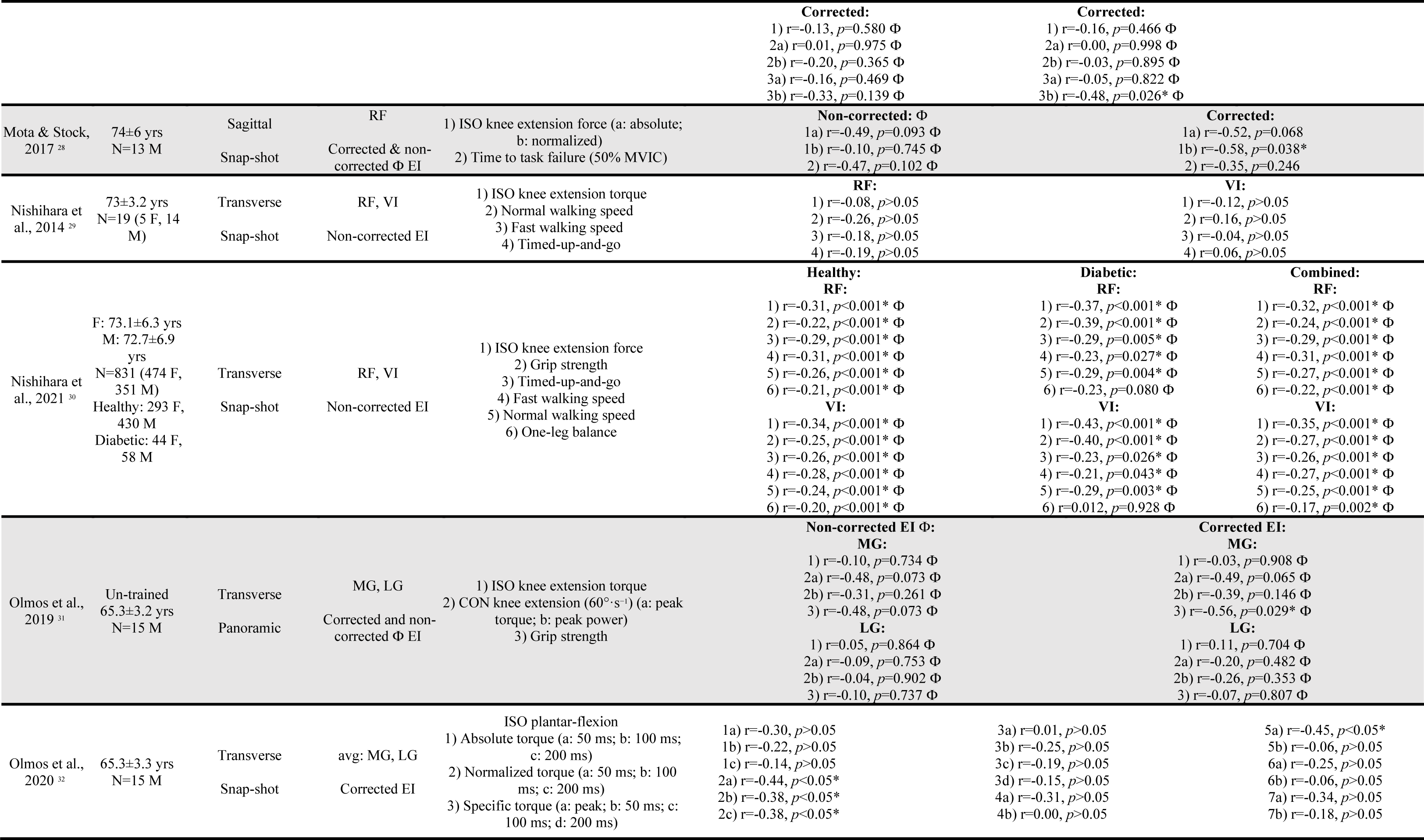
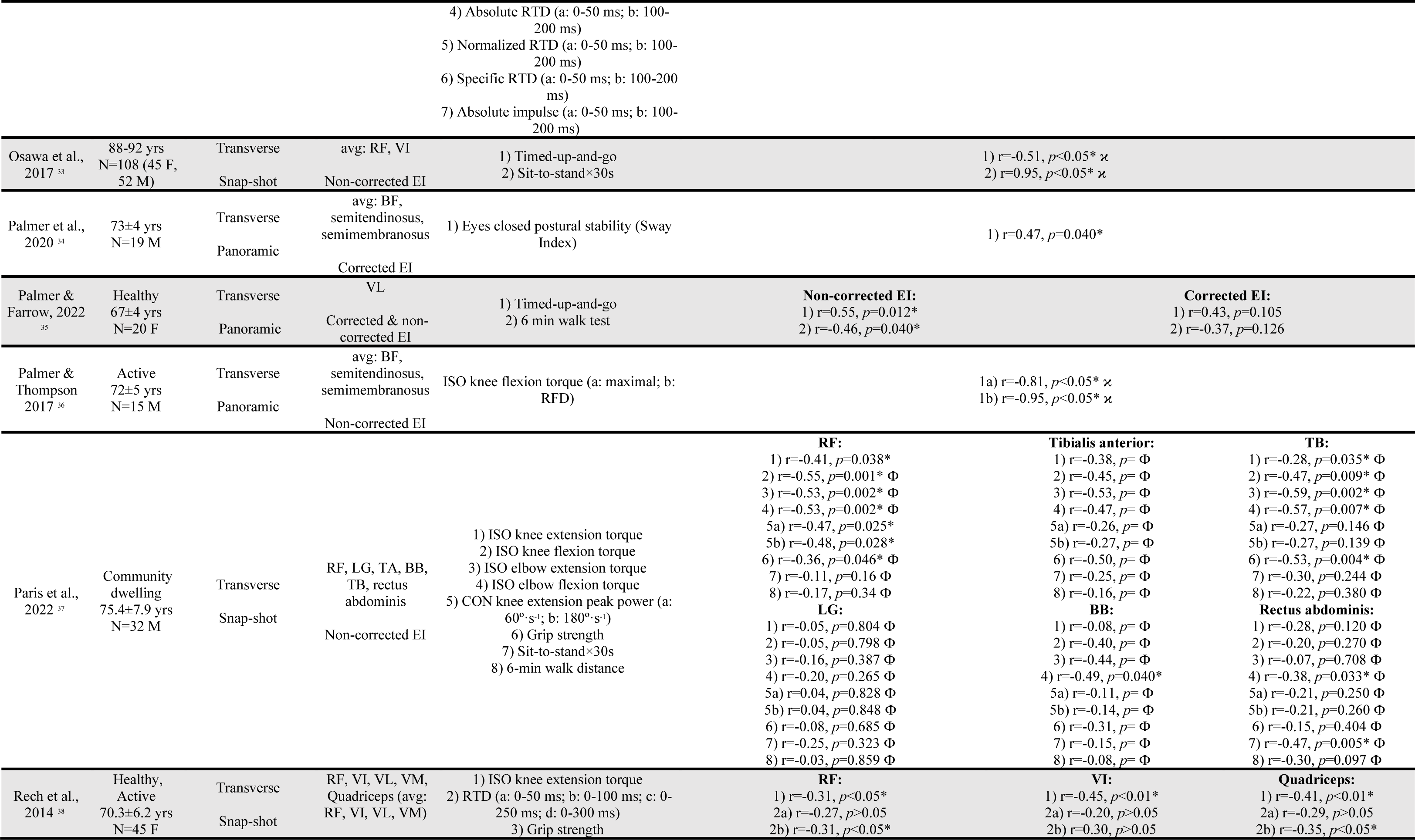
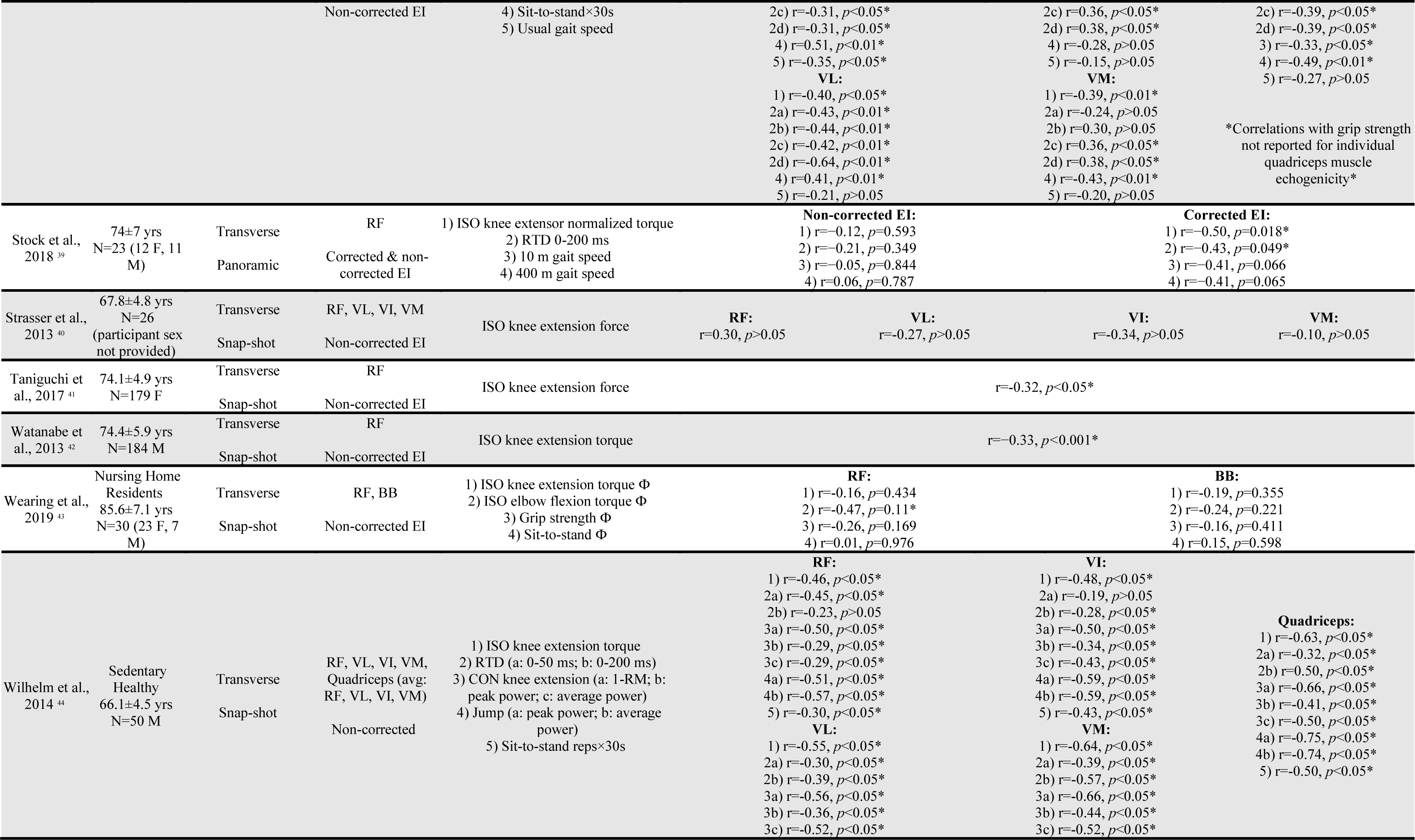
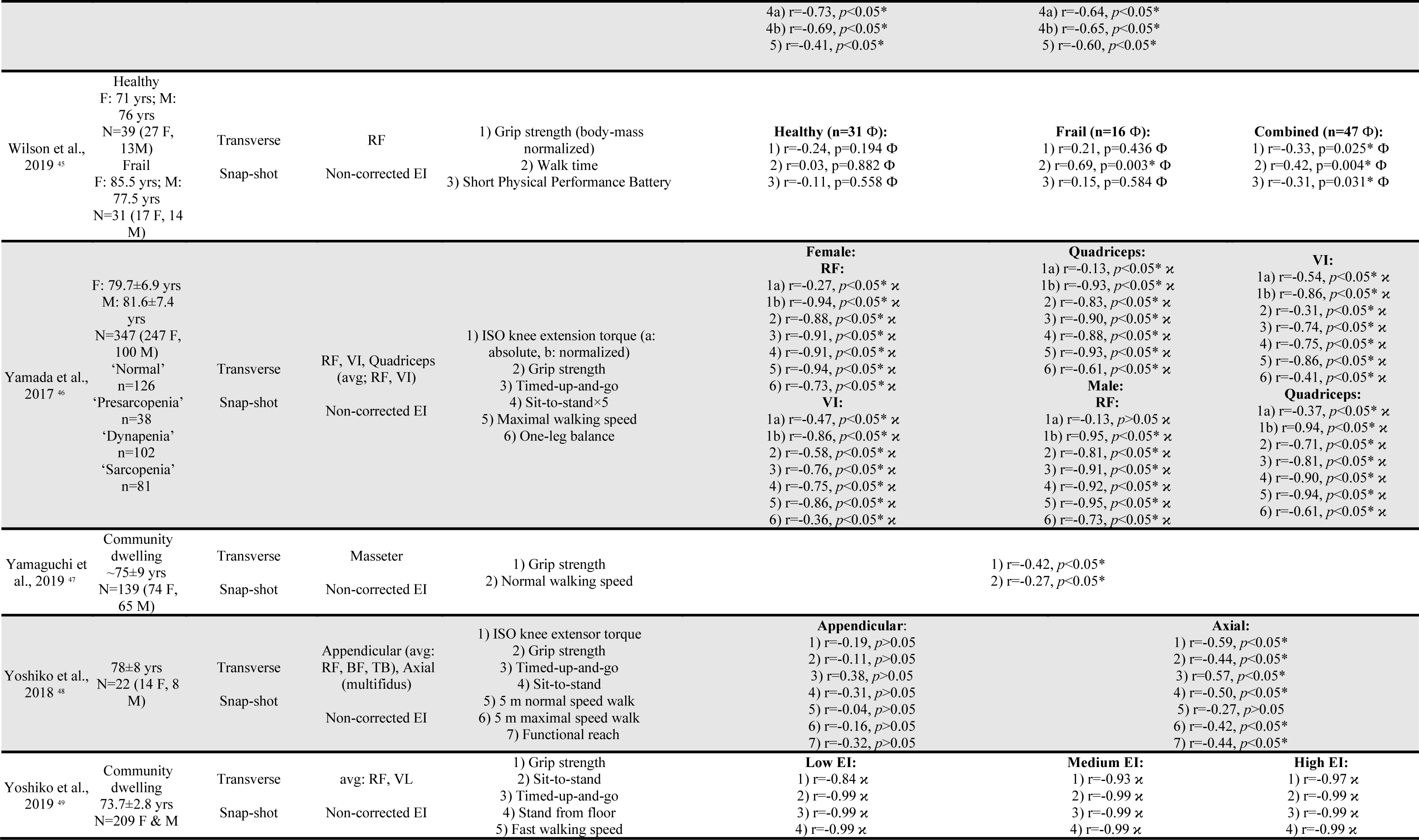
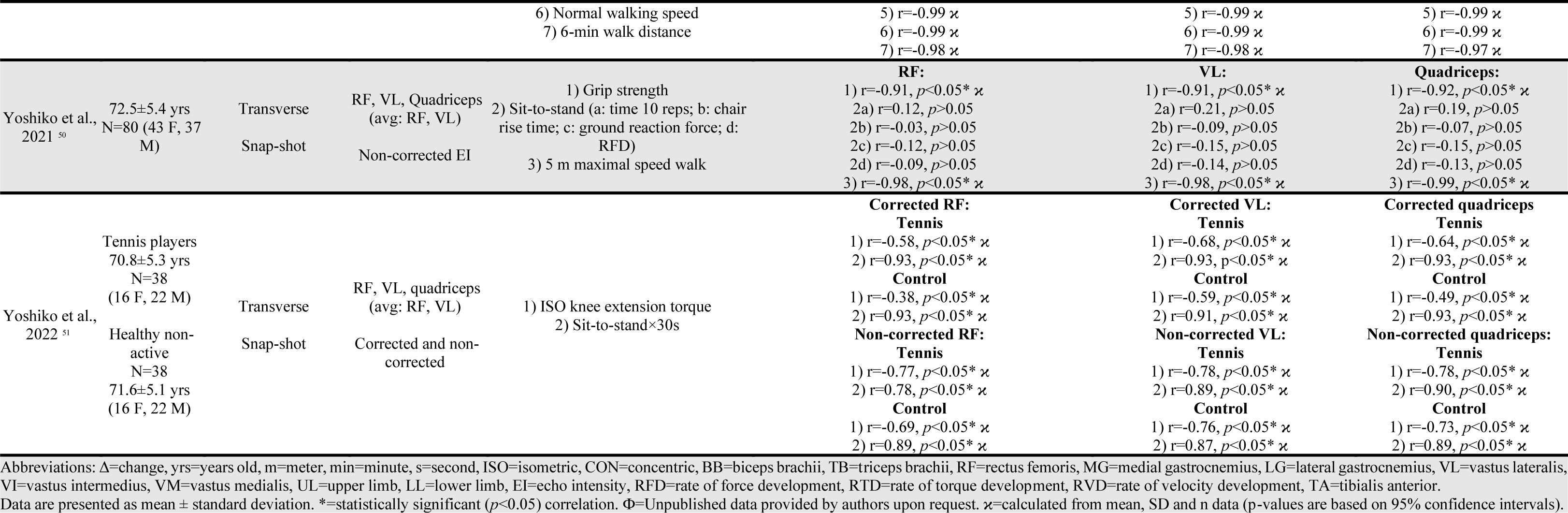
Summary of studies reporting correlations between skeletal muscle echogenicity and functional measures in older (Mean ≥ 63 years) adults.

Scans were collected in the transverse plane in 46 studies,^7,12,14–17,19–24,26–30,32–36,38–44,46–62^ with four obtaining sagittal plane images,^11,18,37,44^ one study utilized both transverse and sagittal scanning planes.^44^ Snap-shot images were collected in 39 studies,^7,11,12,14,17,18,20–24,26–29,32,33,35–42,44– 47,49,53,55–62^ with 11 using panoramic/extended field-of-view technologies.^15,16,19,30,34,43,48,50–52,54^ The scanning plane and scanning method could not be deduced in a single study.^31^ Non-corrected echogenicity was used in 49 studies,^7,11,12,14–20,22–24,26–49,51–62^ while subcutaneous fat-correction was reported or obtained in 12 studies.^15,16,18,19,21,30,40,48,50,51,54,59^ Both non-corrected and corrected values and/or correlations were found or obtained from 10 studies.^15,16,18,19,30,40,48,51,54,59^

The rectus femoris was the most common (n=33) isolated muscle included in _correlations,^7,11,12,14,17,18,20,22–24,28,29,31–37,39–41,43,46,47,53–57,59–62^ followed by the vastus lateralis (n=11),^12,28,29,36,38,39,51,53,56,59,62^ medial gastrocnemius (n=7),^15,16,32,33,36,40,48^ vastus intermedius_ (n=7),^12,20,41,47,53,56,57^ lateral gastrocnemius (n=4),^15,22,40,48^ biceps brachii (n=4),^22,23,32,33^ vastus medialis (n=3),^12,53,56^ tibialis anterior (n=3),^22,36,42^ and triceps brachii (n=3).^22,32,33^ Single studies reported correlations with the multifidus,^44^ erector spinae,^44^ gluteus medius,^38^ masseter,^58^ psoas major,^44^ or rectus abdominis^22^ muscles. Several studies also pooled multiple muscles including all four quadriceps muscles (n=4),^27,41,53,56^ rectus femoris and vastus lateralis (n=7),^28–30,57,59,61,62^ rectus femoris and vastus intermedius (n=2),^49,57^ biceps brachii and the wrist flexors (n=1),^45^ lateral gastrocnemius and soleus (n=1),^26^ rectus femoris, biceps femoris and triceps brachii (n=1),^60^ rectus femoris, vastus intermedius, medial gastrocnemius and lateral gastrocnemius (n=1),^19^ and rectus femoris, vastus lateralis, tibialis anterior and medial gastrocnemius (n=1).^45^

The most common functional test was peak force or torque during isometric knee extension (n=22)^7,12,14,17,18,20,22,23,27,31,35,39,43,47,48,53–57,59,60^ or flexion (n=2),^22,52^ followed by measures of grip strength (n=20),^11,14,20,22–24,28,32,33,37,40,45,46,48,53,57,58,60–62^ walking speed (n=16),^14,17,20,32,33,36,40,44,45,47,53,54,57,58,60,61^ sit-to-stand (n=16),^22,23,28,29,38,40,41,43,49,53,56,57,59–62^ timed up-and-go (n=10),^17,20,38,45,47,49,51,57,60,61^ concentric knee extension force or power, (n=8)^11,22,30,31,39,45,48,56^ isometric or concentric plantar-flexion (n=5) strength, power or velocity, ^15,16,19,26,48^ single leg balance or posture (n=5),^17,20,38,50,57^ isometric elbow flexion (n=2)^22,23^ or extension (n=1),^22^ sit-up (n=2),^28,29^ supine to stand (n=2),^28,29^ ‘functional reach’ score (n=2),^40,60^ or short physical performance battery (n=2).^24,45^ Single studies reported correlations between echogenicity and the ‘functional independence measure gait score’,^27^ heel-rise repetitions,^40^ dorsi-flexion,^42^ or hip flexion force,^34^ jump height or power,^56^ knee extension fatigue,^30^ stand from floor,^61^ or the Tinetti performance test.^45^ Ten studies included at least one measure of rapid force/torque/velocity production.^16,19,21,34,39,52–54,56,62^

### 3.2. Knee extension strength

Twenty studies, including 2924 participants, reported or later provided correlations between quadriceps echogenicity and maximal isometric or concentric knee extension force or torque.^7,12,17,18,20,22,23,23,27,30,31,35,37,39,43,45,47,54–56^ Meta-analysis determined a moderate (*r*=−0.36 [95%CI: −0.38 to −0.32], *p*<0.001) negative correlation between quadriceps echogenicity and knee extension strength in older adults (**Figure 2A**). Sub-group analyses revealed no significant differences between correlations between knee extension strength and individual quadriceps muscles (**Figure 2B**). However, despite relatively few studies for comparison, the vastus medialis muscle holds larger correlations (*r*=−0.41 [95%CI: −0.65 to −0.10], *p*=0.01) when compared to vastus intermedius (*r*=−0.35 [95%CI: −0.41 to −0.30], *p*<0.001), vastus lateralis (*r*=−0.30 [95%CI: −0.55 to −0.02], *p*=0.04), or rectus femoris (*r*=−0.34 [95%CI: −0.37 to −0.30], *p*<0.001) muscles. Interestingly, combining the rectus femoris with vastus lateralis echogenicity (*r*=−0.60 [95%CI: - 0.77 to −0.36], *p*<0.001) outperformed combining all four quadriceps muscles (*r*=−0.48 [95%CI: - 0.72 to −0.13], *p*=0.01). Additionally, the rectus femoris and vastus intermedius combination (Fisher’s Z lower 95%CI: −0.38) was superior to rectus femoris in isolation (Fisher’s Z upper 95%CI: −0.39).

**Figure 2.**
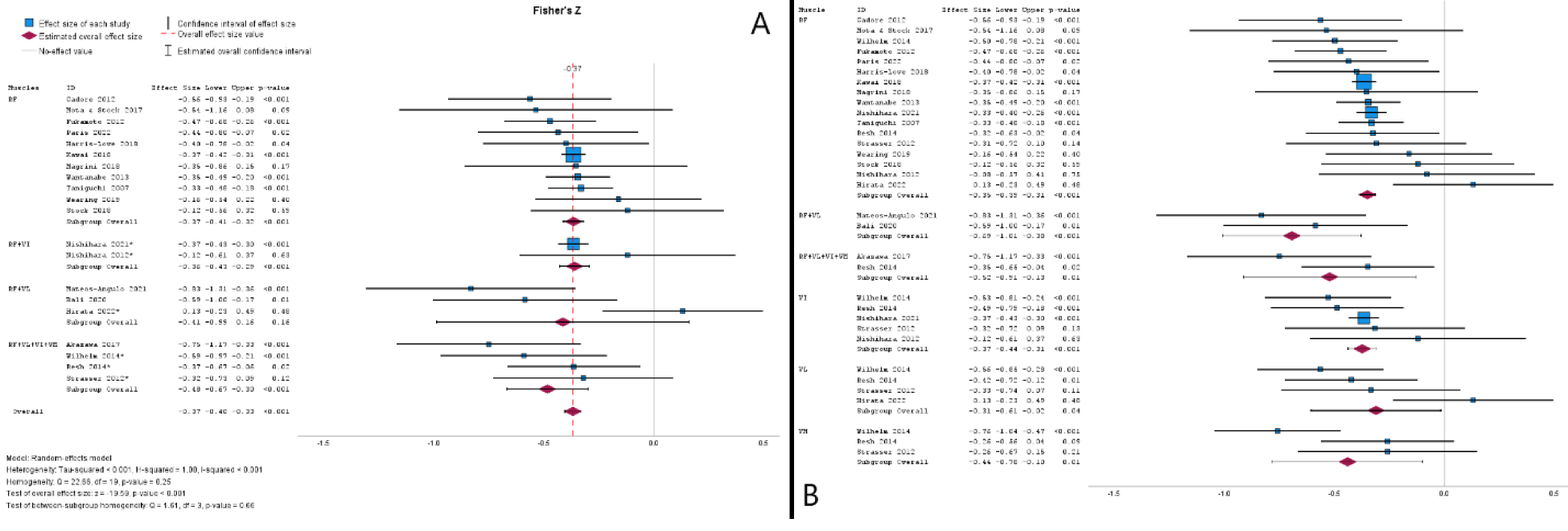
Meta-analytical forest plot of Fisher’s Z correlations between quadriceps echogenicity and maximal isometric or concentric force or torque (**panel A**) with sub-group analysis for individual quadriceps muscles (**panel B**). *=studies where multiple muscles and correlations were averaged.

### 3.3. Grip strength

Sixteen studies, including 1912 participants, reported or provided correlations between muscle echogenicity and grip strength.^11,14,20,22–24,28,32,33,37,40,45,46,48,53,58^ Meta-analysis determined a moderate (*r*=−0.31 [95%CI: −0.37 to −0.24], *p*<0.001) negative correlation between echogenicity and grip strength (**Figure 3**). No significant differences were found when comparing correlations between different individual muscles via sub-group analyses.

**Figure 3.**
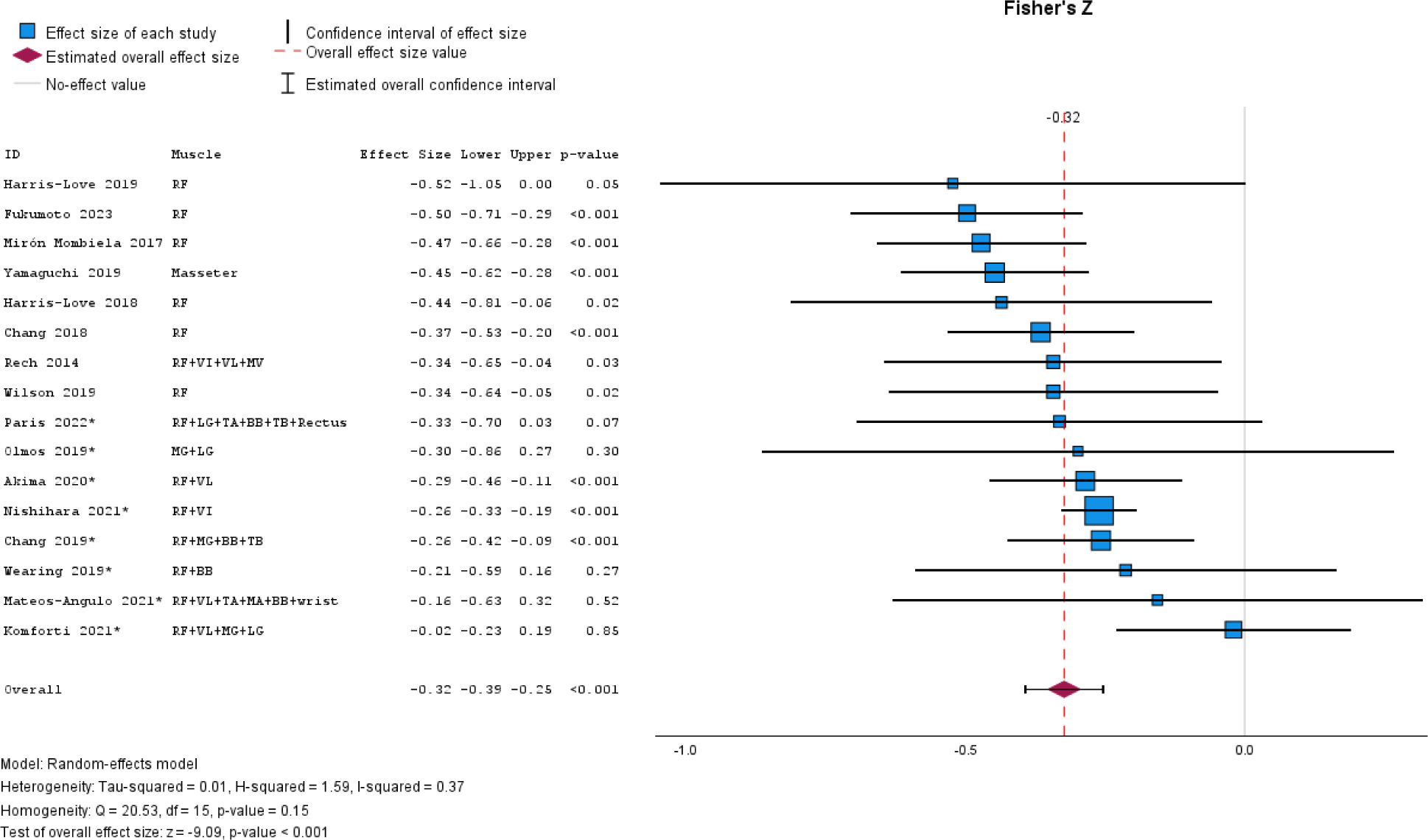
Meta-analytical forest plot of Fisher’s Z correlations between echogenicity and grip strength. *=studies where multiple muscles and correlations were averaged.

### 3.4. Walking speed

Within study comparison demonstrated that preferred or normal walking speed (*r*=−0.11. range: −0.46 to 0.17) held smaller correlations (*p*=0.015) with muscle echogenicity when compared to maximal walking speed (r=−0.17, range: −0.51 to 0.15).^20,36,40,44,47,60,61^ Therefore, the meta-analysis only included maximal walking speed where possible. The meta-analysis included 17 studies, including 2972 participants.^14,17,20,22,24,28,29,33,36,40,44,45,51,53,54,58,60^ Meta-analysis determined a small (*r*=−0.23 [95%CI: −0.29 to −0.16], *p*<0.001) negative correlation between echogenicity and walking speed (**Figure 4**). Not enough between-study consistency existed to examine sub-analysis of individual muscle groups.

**Figure 4.**
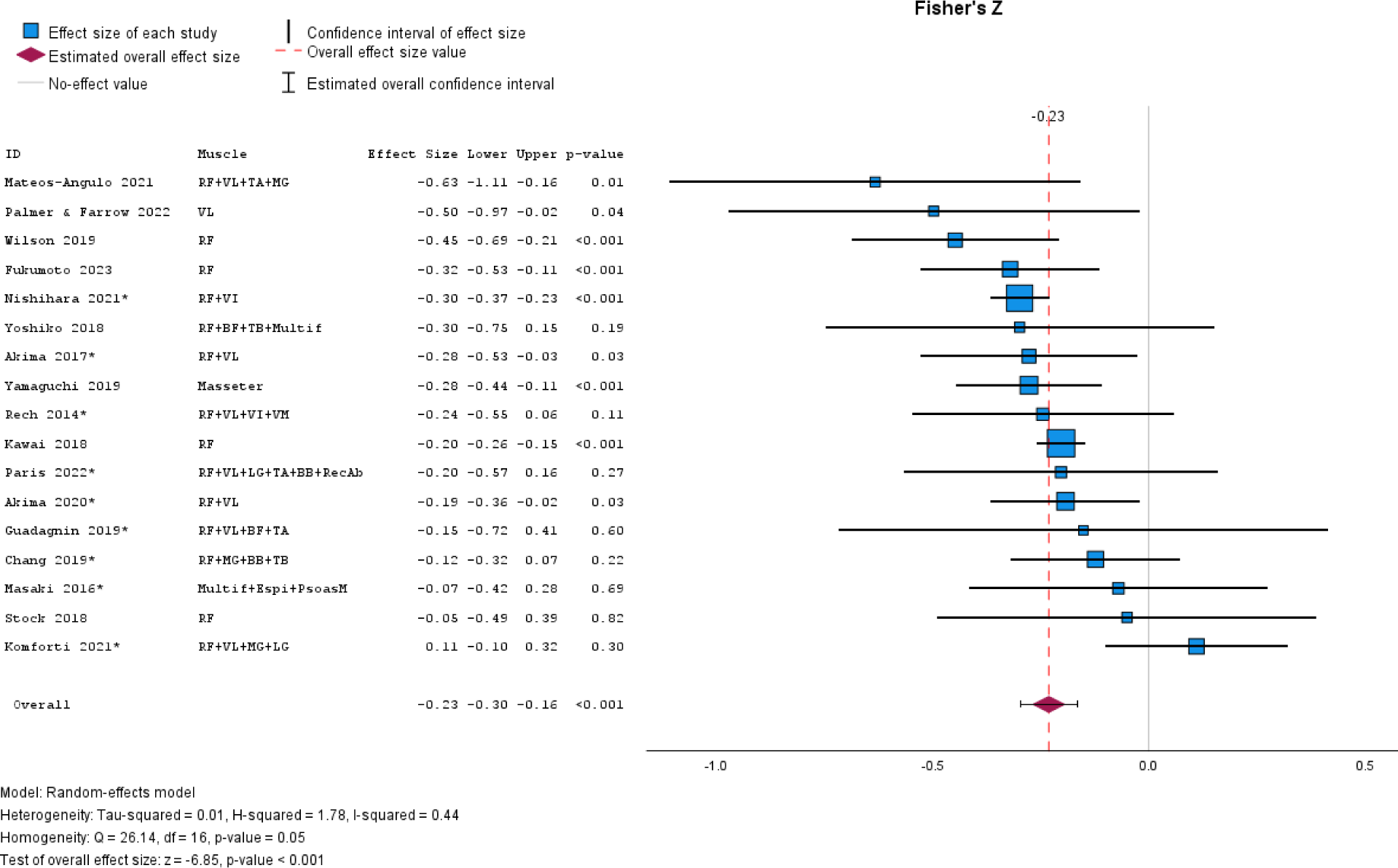
Meta-analytical forest plot of Fisher’s Z correlations between echogenicity and walking speed. *=studies where multiple muscles and correlations were averaged.

### 3.5. Sit-to-stand

A total of 12 studies with 666 elderly participants were included in the meta-analysis determining a moderate (*r*=−0.34 [95%CI: −0.44 to −0.23], *p*<0.001) negative correlation between echogenicity and sit-to-stand performance (**Figure 5A**).^22,23,28,29,38,40,41,43,53,56,60,62^ When examining individual muscles (**Figure 5B**), the combination of all four quadriceps muscles (*r*=−0.61 [95%CI: −0.74 to −0.43], *p*<0.001) appears to have superior correlations when compared to rectus femoris (*r*=−0.25 [95%CI: −0.37 to −0.14], *p*<0.001), vastus lateralis (*r*=−0.38 [95%CI: −0.51 to −0.21], *p*<0.001), rectus femoris with vastus lateralis (*r*=−0.28 [95%CI: −0.45 to −0.10], *p*=0.03), or vastus intermedius (*r*=−0.45 [95%CI: −0.70 to −0.10], *p*<0.001). While only incorporating two studies, vastus medialis (*r*=−0.55 [95%CI: −0.73 to −0.37], *p*<0.001) outperformed all other individual quadriceps muscles when correlating to sit-to-stand performance.

**Figure 5.**
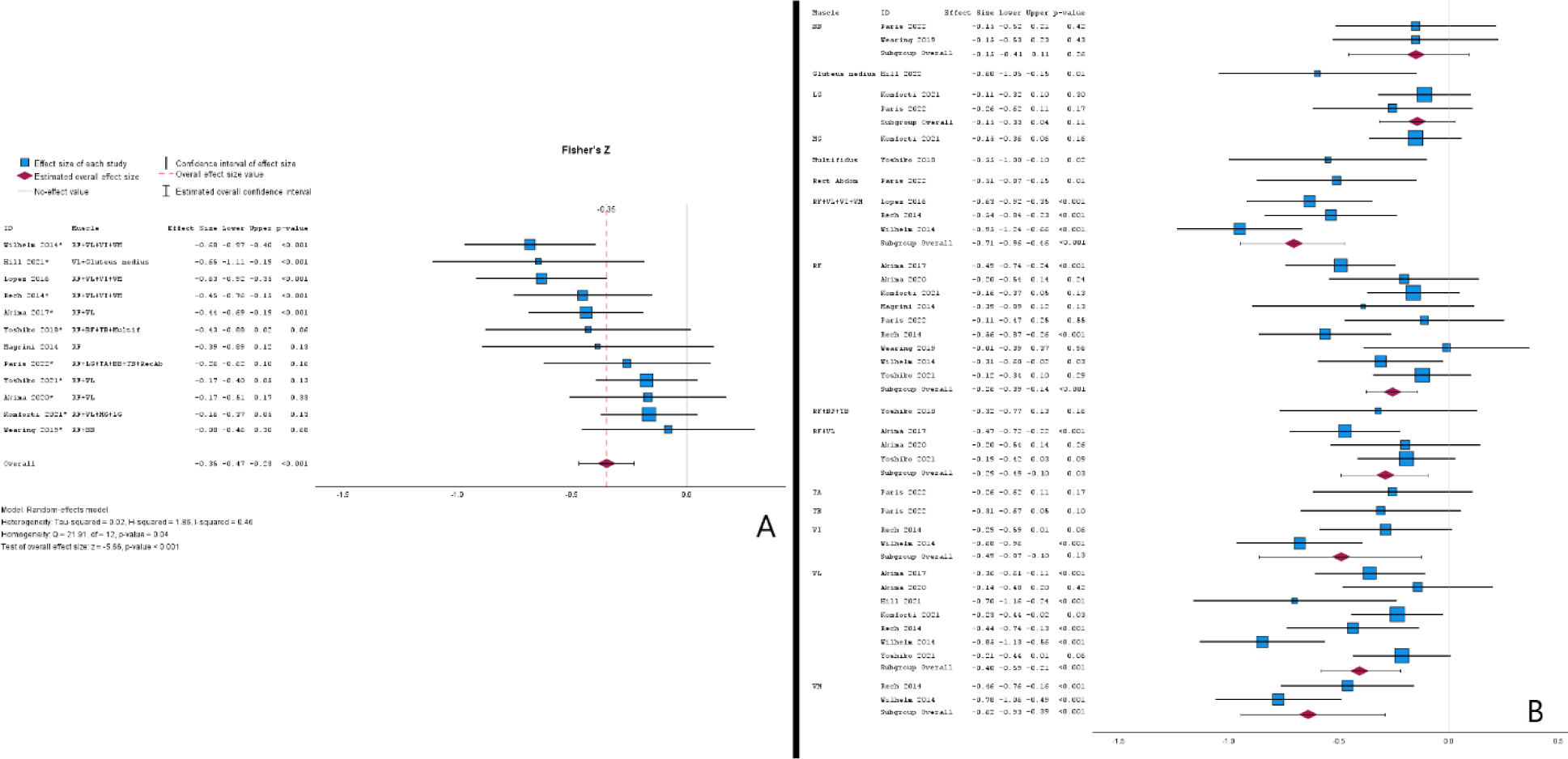
Meta-analytical forest plot of Fisher’s Z correlations between quadriceps echogenicity and sit-to-stand performance (**panel A**) with sub-group analysis for individual quadriceps muscles (**panel B**). *=studies where multiple muscles and correlations were averaged.

### 3.6. Timed up-and-go

Seven studies, incorporating 2172 participants, were included in a meta-analysis. The meta-analysis determined a small (*r*=−0.26 [95%CI: −0.35 to −0.18], *p*<0.001) negative correlation between echogenicity and timed up-and-go performance (**Figure 6**).^17,20,38,45,47,51,60^ Sub-group analyses could not be confidently performed. However, it seems plausible that vastus lateralis (*r*=−0.55 to −0.51) echogenicity is a better predictor of timed up-and-go performance than the rectus femoris (*r*=−0.29 to −0.19) or vastus intermedius (*r*=−0.26 to 0.06). Interestingly, the largest single correlations with timed up-and-go performance were multifidus (*r*=−0.57) and gluteus medius (*r*=−0.49) echogenicity.

**Figure 6.**
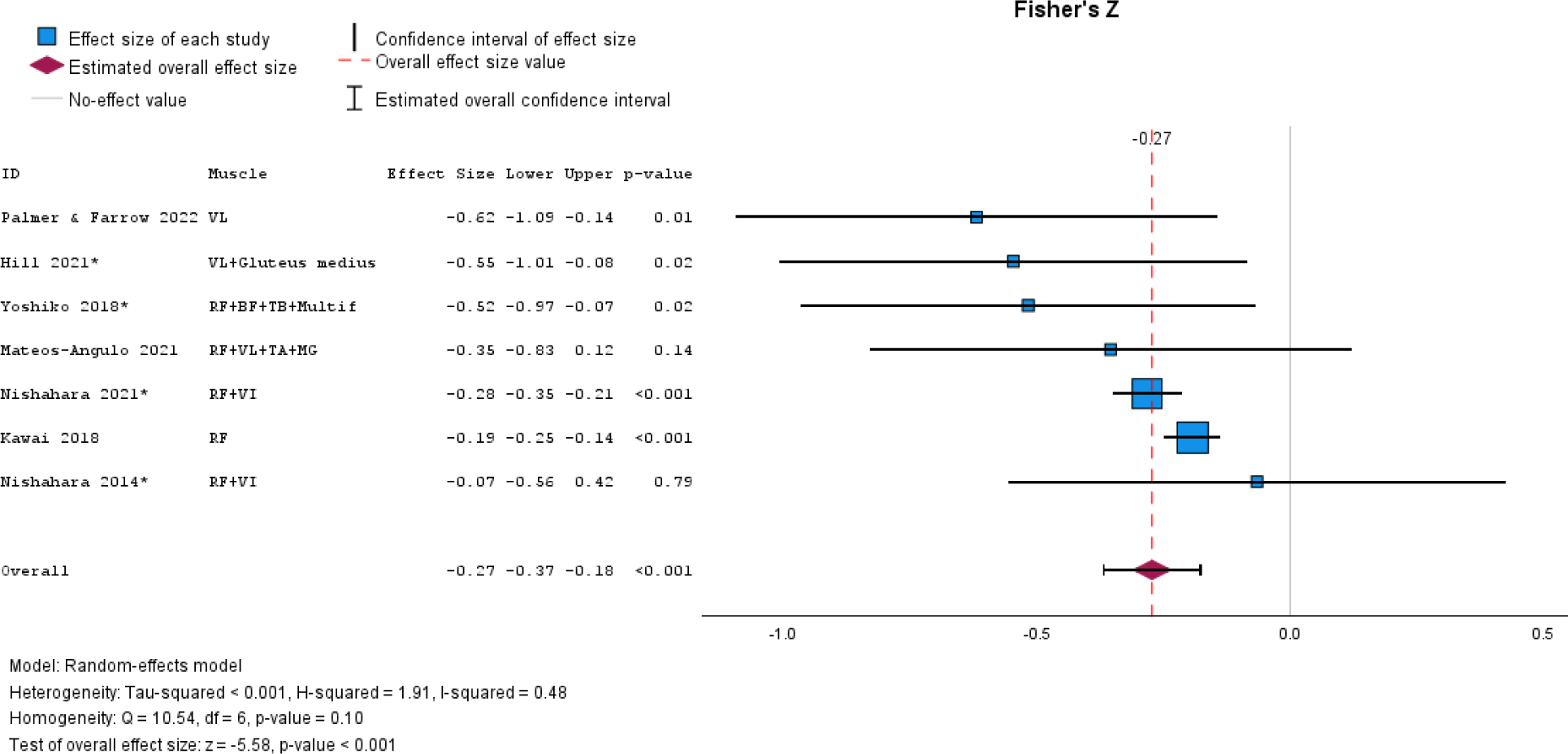
Meta-analytical forest plot of Fisher’s Z correlations between echogenicity and timed up-and-go performance. *=studies where multiple muscles and correlations were averaged.

### 3.7. Rapid force production

Ten studies report measures of rapid force,^16,52,62^ torque,^21,34,53,54,56^ or velocity^19,39^ development. Correlations with echogenicity were generally small and highly variable (*r*=−0.15; range: −0.42 to 0.25). Methods were too variable for meta-analyses to be confidently performed.

### 3.8. Additional considerations

Other physical performance measures (e.g., sit-up, functional batteries, balance tasks) did not exist in large enough quantities to warrant data pooling. However, correlations were trivial to large (*r*=−0.58 to 0.23) and highly variable. Funnel plots illustrate minimal risk of publication bias **Figure 7**.

**Figure 7.**
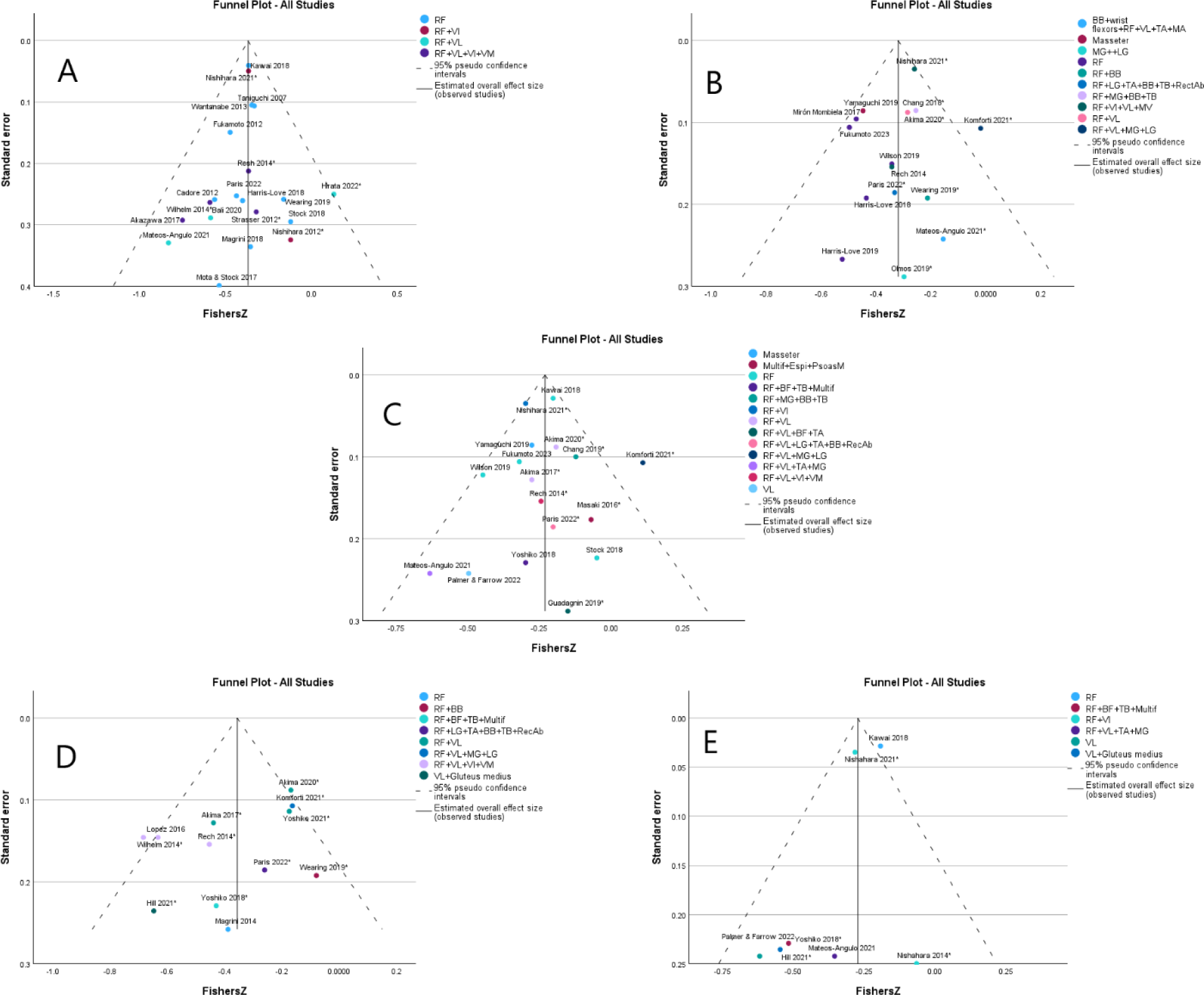
Funnel plots representing echogenicity correlations with knee extension strength (panel A), grip strength (panel B), walking speed (panel C), sit-to-stand (panel D), and timed up-and-go (panel E) performances, respectively

## 4. Discussion

Early identification of skeletal muscle weakness or dysfunction is imperative for early diagnosis and treatment for older individuals. Ultrasound-derived echogenicity is one potential method to detect and track changes in muscle composition and loss of function. Therefore, this systematic review with meta-analyses aimed to compile available research that evaluates the relationships between muscle echogenicity and measures of muscle functional assessments in older adults. The meta-analyses, consisting of 7-20 studies, consistently found moderate, negative relationships, demonstrating higher measures of echogenicity (lower muscle quality) associated with lower physical function. However, results were not uniform across all studies.

This study’s primary and largest meta-analysis examined the relationships between quadriceps muscle echogenicity and knee extension strength. This pooled analysis included 20 studies and 2924 older adults^7,12,17,18,20,22,23,23,27,30,31,35,37,39,43,45,47,54–56^ and demonstrated a modest, negative relationship, with relatively narrow confidence intervals (*r*=−0.36 [95%CI: −0.38, −0.32], *p*<0.001). Quadriceps strength has demonstrated moderate to strong relationships to functional tasks,^23^ with lower quadriceps strength increasing the odds (OR=3 [95%CI, 1.78 to 5.05]) of falls within older adults.^64^ Further, quadriceps strength relates to mortality, demonstrating a high utility for patient prognoses.^65^ The current review demonstrated that skeletal muscle quality quantified through ultrasound echogenicity might explain a relatively small variance of knee extensor torque. Prior research has found that echogenicity and muscle volume predict greater variances of knee extensor torque compared to muscle volume alone,^66^ which may suggest that morphological components of skeletal tissue contribute to the muscle’s functionality. The small to moderate pooled estimates within the current review may support echogenicity as a supplementary outcome to muscle volume in predicting strength or function. However, quadriceps echogenicity may be an appropriate surrogate to quantify knee extensor torque and total-body physical function as a stand-alone measure. The consistent findings of negative relationships may propose the utility of such measures within a screening protocol in a battery of other assessments. Greater intramuscular adipose tissue is associated with an 8% risk of mortality^67^ and has thus been suggested as a time-efficient and noninvasive sarcopenia screen.^68^ Finally, it should be noted that sub-analysis, which including a limited number of studies excluding the rectus femoris, demonstrated relatively minimal differences in correlations with knee extensor strength over different quadriceps muscles. However, the studies combining multiple quadriceps muscles (rectus femoris+vastus lateralis; rectus femoris+vastus lateralis+vastus intermedius+vastus medialis) demonstrated the strongest correlations.

Like quadriceps strength, the meta-analysis of grip strength demonstrated a modest negative correlation (*r*=−0.31 [95%CI: −0.37 to −0.24], *p*<0.001) with echogenicity. When examining non-meta data, lower tissue quality in other muscle groups was also negatively related to knee extensor torque. For example, the multifidous and gastrocnemius echogenicity demonstrated weak to moderate negative relationships to knee extensor torque.^60^ Prior research has shown lower muscle quality present across multiple muscle groups with older individuals – suggesting global changes in muscle morphology, a finding supported by the present meta-analyses. Mateos-Angulo et al.^45^ found that the scanning location for muscle quality is important in collecting different strength measures. Muscle echogenicity contained in the upper extremity (biceps brachii and superficial wrist flexors) and lower extremity (rectus femoris, vastus lateralis, medial gastrocnemius, and tibialis anterior) were found to have moderate relationships to knee extensor torque, but not to hand grip strength.^45^ Hand-grip strength is a commonly collected measure within older adults to screen for sarcopenia.^1,69^ The current review demonstrates its relationship to skeletal muscle quality to be inconsistent across and within muscle groups (*r*=−0.52 to −0.02). Chang et al.^32^ found significant, negative relationships to hand grip strength when muscle quality was assessed within the biceps brachii, triceps brachii, and rectus femoris, but not to the medial gastrocnemius. Yoshiko et al.^60^ averaged four scanning locations (rectus femoris, biceps femoris, and triceps brachii) and found no significant relationships to hand-grip strength.

Relationships to dynamic performance (walking speed: *r*=−0.23 [95%CI: −0.29 to −0.16], *p*<0.001; sit-to-stand: *r*=−0.34 [95%CI: −0.44 to −0.23], *p*<0.001; timed up-and-go: *r*=−0.26 [95%CI: −0.35 to −0.18], *p*<0.001) were similar, but slightly weaker and less homogenous than quadriceps strength. Intermuscular adipose fat has been demonstrated to be associated with annual declines in gait speed, a relationship independent from total body adiposity in men.^70^ Maximal gait speed in older adults are influenced by numerous neuromuscular factors, which may limit the predictability of such task from quality assessment of a single muscle.^71^ Also, the number of individual muscles needed to be recruited and activated for walking tasks may explain the inconsistencies or lack of relationships between a single muscle group and gait speed. Nevertheless, the relatively similar between-variable pooled correlations, combined with the sub-analyses, suggest that muscle echogenicity is systemic; thus, poor muscle quality in one muscle is likely representative of total-body skeletal muscle composition.

In addition to strength, the current review extracted ten studies that assessed the relationship between muscle echogenicity and rapid force, torque, or velocity production.^16,19,21,34,39,52–54,56,62^ Rate of force/torque/velocity development quantifies the time frame at which these qualities are expressed and often hold stronger relationships to sport-specific and functional daily tasks than strength alone.^72,73^ Rapid strength expression is limited in older, active men compared to a younger cohort.^74^ Negative, but highly variable relationships were observed between muscle echogenicity and rapid force expression, though the methods by which rapid force expression was measured may influence these clinical relationships. Olmos et al.^21^ assessed the rate of torque development in terms of absolute (non-normalized), normalized (normalized to peak torque), or specific (normalized to muscle cross-sectional area) and only found weak, negative relationships to exist between gastrocnemius echogenicity and plantar-flexion rate of torque development when normalized to peak torque.^21^ Additionally, the time domain in calculating rapid force expression may contribute to the relationships to clinical outcomes. Studies extracted in this review calculated rapid force from 0-50 ms to 0-200 ms, demonstrating weak to moderate relationships to echogenicity with each.^21,56^

### 4.1. Limitations and future research directions

While a substantial number of studies, participants, muscles, functional tests, and analyses were examined, this systematic review with meta-analyses has limitations. While this review focused on older adults, ultrasonic muscle quality and composition estimations are potentially valuable to various populations, including development through adulthood, athletes, and people with neuromuscular conditions. Due to general differences in muscle fiber type and body fat amount and distribution, the effects of sex on the relationships between skeletal muscle echogenicity is another exciting topic, especially considering the lack of studies reporting correlations with subcutaneous fat thickness corrected values despite correction resulting in greater inter-session reliability.^75^ While one of the strengths of meta-analysis is to balance out studies with extreme findings, it should be noted that Kawai et al.,^17^ (N=1239), and Nishihara et al.,^20^ (N=831) included the largest sample sizes by a substantial magnitude. Thus, these two studies affected the knee extension strength, grip strength (only Kawai et al., 2018), walking speed, and timed up-and-go analyses more than others. However, these two studies were never ‘outliers’ compared to the other relevant studies. Finally, this review included only cross-sectional correlational studies. Therefore, future meta-analytical reviews may wish to examine the relationships between longitudinal changes in echogenicity and physical function.

## 5. Conclusions

The results of this systematic review with meta-analyses demonstrate a consistent yet modest association between skeletal muscle echogenicity and physical function in ageing adults. Additionally, sub-analyses show minimal between-muscle differences in correlations between echogenicity and physical function, suggesting that ultrasound-estimated muscle quality and composition are systemic. However, including multiple muscles tends to improve the predictive ability of muscle strength measures. Researchers and practitioners should, therefore, understand echogenicity screenings for muscle-related issues should be supplemented with additional assessments, such as muscle thickness. Additionally, while researchers could consider combining multiple muscles to improve correlational strength, time-poor practitioners can choose to scan a single, easily accessible muscle to estimate total body muscle composition.

## Acknowledgements

We thank Dr’s Daisy Wilson, Eric Ryan, Garret Hester, Gena Gerstner, Hisashi Kawai, Jacob Mota, Julia Wearing, Matt Stock, Michael Paris, and Yoshihiro Fukumoto for providing additional findings from their publications.

## Authors’ contributions

MHL provided salary support for all other authors. MHL and SGB conceived the initial topic. SGB, KLB and DJO performed literature searches and study evaluations. DJO, SGB, and KLB extracted the study data. DJO performed the statistical analyses and created the figures. DJO and SGB wrote the introduction and discussion sections, while DJO wrote the abstract, methods, and results. All authors edited and approved the submitted manuscript.

## Data Availability

This is a meta-analysis, therefore, all underlying data is already available.

